# Context-specific impact of antimicrobial stewardship on antibiotic use and antibiotic resistance in hospitals in a lower-middle income country - results from an implementation study with a controlled interrupted time series design in Vietnam

**DOI:** 10.1101/2025.08.25.25334341

**Authors:** Le Quynh Trang, Vu Tien Viet Dung, Le Minh Quang, Nguyen Thi Thu Huyen, Vu Hai Vinh, Chau Minh Duc, Vo Thi Hoang Dung Em, Nguyen Thi Cam Tu, Truong Anh Quan, Nguyen Hong Khanh, Le Nguyen Minh Hoa, Thomas Kesteman, Elizabeth Dodds Ashley, Deverick J. Anderson, Hugo C Turner, Pham Ngoc Thach, Ben S Cooper, Marc Choisy, H Rogier van Doorn, Vu Thi Lan Huong

## Abstract

**Introduction:** High-quality evidence regarding the impact of antimicrobial stewardship (AMS) is limited in Asia. In this study, we aimed to determine the effects of a pharmacist-led prospective audit with feedback intervention, as part of an AMS programme following national guidelines, in two provincial-level general hospitals in Vietnam, a lower-middle-income country.

**Methods:** We performed controlled interrupted time-series analyses to evaluate the impact of an AMS intervention on antibiotic use in days of therapy per 1000 patient-days, antibiotic non-susceptibility percentage and patient outcomes. In each hospital, four wards received the intervention and four wards acted as controls. Pre-intervention periods began in January 2019 and continued to May 2020 (Hospital 1) and July 2020 (Hospital 2), followed by a 12-month post-intervention period.

**Results:** In Hospital 1, the intervention was associated with a reduction in the level of antibiotic use (95.9, 95% CI [10.9, 180.8]), although there was no evidence for a change in trend (0.9[−3.6, 5.4]). In contrast, in Hospital 2, there was no evidence for a change in either level (6.3[−83.7, 96.3]) or trend (−2.1[−4.8, 0.6]). In Hospital 1, we observed a decreasing trend in antibiotic non-susceptibility among hospital-acquired Escherichia coli to aminoglycosides (odds ratio: 0.87[0.78, 0.97]), but increasing for Pseudomonas aeruginosa to carbapenems (1.11[1.00, 1.22]) and Acinetobacter spp. to aminoglycosides (1.07[1.00, 1.27]). In Hospital 2, evidence indicated decreasing trends in Acinetobacter spp. to carbapenems (0.96[0.88, 1.00]), ciprofloxacin (0.93[0.85, 1.00]), and piperacillin-tazobactam (0.94[0.78, 1.00]), but increasing for P. aeruginosa to aminoglycosides (1.07[1.00, 1.20], ciprofloxacin (1.45[1.18, 1.77]), and ceftazidime (1.03[1.00, 1.19]). We did not find evidence that the intervention was associated with changes in mortality or hospitalisation costs.

**Conclusion:** The impact of AMS varied between the two hospitals, highlighting context-specific implementation challenges and the necessity to monitor changes in antibiotic resistance over time to tailor interventions that respond to local resistance epidemiology.

**WHAT IS ALREADY KNOWN ON THIS TOPIC:**

- Previous systematic reviews and recent interrupted time series (ITS) studies have shown large variations in the impact of antimicrobial stewardship (AMS) on hospital antibiotic use and resistance. Few previous ITS studies of AMS have used a control in their analysis.

**WHAT THIS STUDY ADDS:**

- Using a strong quasi-experimental study design with a control group, we generated empirical evidence on the multi-faceted effects of an AMS intervention on antibiotic use and resistance in hospitals in a middle-income country in Asia.
- The implementation of prospective audit and feedback in the context of established AMS programmes following the national guidelines had different effects on total antibiotic use and antibiotic non-susceptibility proportions among common hospital-acquired pathogens found in Vietnam.

**HOW THIS STUDY MIGHT AFFECT RESEARCH, PRACTICE OR POLICY:**

- This study contributes evidence from a strong study design on the effects of AMS implementation in hospitals in a middle-income country in Asia, which can inform future AMS programmes in similar settings.
- It highlights the need to monitor the emergence and spread of antibiotic resistance in hospital settings in Asia to support development and design of novel interventions for AMS and other programmes to respond to the fast-changing resistance profiles of bacterial pathogens in hospitals.

## Introduction

Widespread use of antibiotics in humans, food production animals and spillover into the environment has accelerated the emergence and transmission of drug-resistant bacteria^1^. Antimicrobial stewardship (AMS) interventions are designed to target inappropriate antibiotic use to reduce selective pressure on resistant bacteria^2^. A recent meta-analysis of the global impact of AMS programmes reported that such interventions were associated with an estimated mean reduction in the proportion of patients receiving antibiotic prescriptions by 10% (95% CI: [4%, 15%]) and a mean rate ratio of 0.72 (95%CI: [0.56, 0.92]) in the consumption rate measured by defined daily doses per 100 patient-days^3^. Importantly, AMS programmes need to monitor the impact on mortality to ensure interventions do not harm patients as well as impact on antibiotic use and resistance. Unfortunately, such evidence for hospital inpatients is currently insufficient^4 5^. A meta-analysis including 221 studies across 34 countries by Davey et al found that mortality risks were similar between intervention and control groups^4^.

The two study designs considered appropriate for evaluating the impact of AMS interventions are randomised controlled trials and quasi-experimental studies (non-randomised controlled trials, controlled before-and-after designs, and interrupted time series)^4^. Recent evidence of the effect of AMS interventions on resistance outcomes comes from studies with weak designs. For example, between 2012 and 2017, only 8 of 26 studies used interrupted time series, and none included a control group in their analysis^5^. In recent studies, only one study in Canada used a control group and demonstrated the impact of a comprehensive AMS programme with a sustained reduction of hospital-acquired antibiotic-resistant organisms^6^. Large heterogeneity in study designs, AMS interventions, and how resistance was evaluated (denominators of outcome measures), along with uncontrolled context-specific confounding factors, have contributed to variations in the reported impact of AMS programmes on resistance^5^.

In this study, we aimed to evaluate the impact of an AMS intervention in two provincial general hospitals (Hospital 1 and Hospital 2) in Vietnam, a lower-middle-income country in Asia. We hypothesised that the AMS intervention would reduce antibiotic use without negatively impacting patient outcomes, and reduce the proportions of patients carrying resistant organisms for clinically important bacterial species. Vietnam developed its first national action plan for controlling antimicrobial resistance and initiated discussions on AMS implementation in a local hospital network in 2013^7^. Since then, several guidelines for antibiotic treatment and AMS implementation in hospitals have been issued and local hospitals have started their AMS programmes following guidelines from the Ministry of Health (MoH) ^8^. We previously reported on our implementation research to assess the feasibility of AMS interventions in these two provincial hospitals in collaboration with the Duke Antimicrobial Stewardship Outreach Network^9^. The theoretical framework for AMS implementation in this research was based on the assumption that hospitals are complex adaptive systems and that AMS teams could leverage their unique characteristics and interconnections to develop a locally feasible and sustainable programme. Prospective audit and feedback (PAF) was chosen as the core AMS intervention implemented at these two hospitals based on evidence from a previous systematic review on effective behaviour change interventions for antibiotic prescribing in hospitals^4^.

## Methods

### Study setting and population

The study was implemented in two provincial hospitals in Vietnam: Hospital 1 (1000 beds) and Hospital 2 (2000 beds). We selected 8/26 clinical wards in Hospital 1 and 8/27 clinical wards in Hospital 2, equally divided between intervention and control groups. Ward selection and assignment were described previously^9^. Briefly, wards were selected based on two criteria: (1) higher-than-average antibiotic use in the hospital based on pharmacy-reported data and (2) willingness of the ward head to participate. All inpatients in the study wards during evaluation periods were included. Allocations of the wards in the two hospitals were similar in terms of clinical specialties, with four intervention versus control ward pairs, as shown in Figure 1. Detailed characteristics of these two hospitals are presented in Table 1S (Supplementary Data).

**Figure 1.**
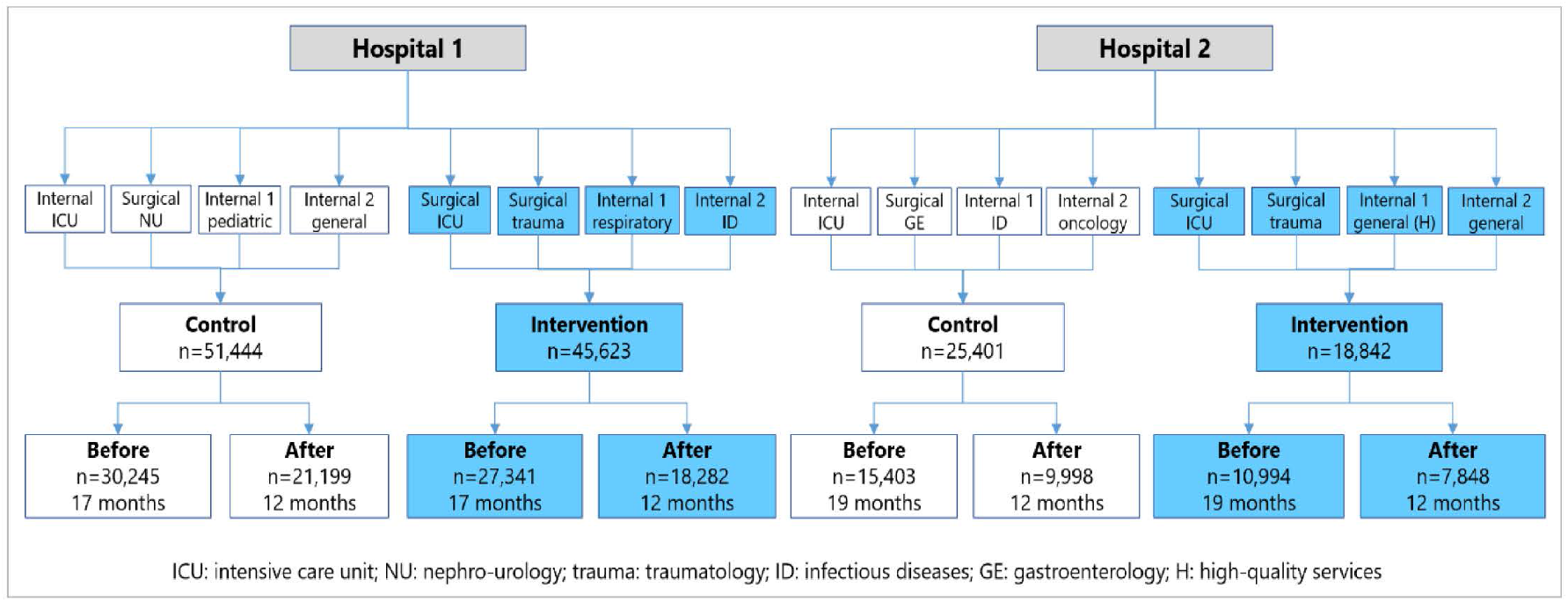
Patient location before and after the AMS implementation start at Hospital 1 (Jan 2019 – May 2021) and Hospital 2 (Jan 2019 – Jul 2021). There were 4 pairs in each Hospital: 1) ICU pair: surgical ICU versus internal ICU (both hospitals); 2) Surgical pair: traumatology versus nephro-urology (Hospital 1) and traumatology versus gastroenterology (Hospital 2); 3) Internal pair 1: respiratory versus paediatrics (Hospital 1); general internal medicine (high-quality services) versus infectious diseases (Hospital 2); and 4) Internal pair 2: infectious diseases versus general internal medicine (Hospital 1); general internal medicine (normal services) versus oncology (Hospital 2). n: number of patient.

Before the intervention, an AMS team was formally established at the two hospitals with the implementation of the pre-authorisation policy (which required doctors to obtain approval from the head of the clinical ward and a director board representative before using antibiotics on the restricted antibiotic list) following the 2016 national AMS guidelines^10^. Infection prevention and control (IPC) guidance from the MoH has been issued since 2009 (18/2009/TT-BYT) with documents and training provided by the MoH to the hospitals since then^11^. The 2016 AMS guidelines included the role of IPC staff in the AMS committee and described the set of responsibilities in implementing protocols for the isolation of patients with multidrug-resistant organisms, along with basic IPC measures such as hand hygiene, use of personal protective equipment, sterilisation of medical equipment, enhanced monitoring, and outbreak investigations. Although there were cases of COVID-19 in 2020 in some specific areas of Vietnam, the COVID-19 pandemic did not affect the two hospitals until late April 2021 in Hospital 1 and July 2021 in Hospital 2, towards the end of the intervention periods^12^.

### Interventions

At the beginning of the project, an AMS team was established to collect baseline data for assessments of needs, gaps, strengths, and weaknesses to inform planning of the intervention^9^. As part of the PAF activity, clinical pharmacists made weekly visits to the intervention wards to review antibiotic prescriptions for patients and provide recommendations for improvement where needed. During this intervention period, both hospitals still maintained the pre-authorisation policy and routine hospital-level IPC activities in all clinical wards as usual, including the study wards. Figure 1S outlines the timeline of project activities implemented before, during, and after the intervention period, when PAF started on 01 June 2020 in Hospital 1 and 29 July 2020 in Hospital 2.

During the one-year intervention period, the PAF activity at Hospital 1 was led by two clinical pharmacists who conducted a total of 1,890 PAF reviews, while Hospital 2 assigned four clinical pharmacists to conducted a total of 1,628 PAF reviews^9^. 82 recommendations were made among the reviews at Hospital 1 (75% were accepted by the treating doctors), and 128 were made at Hospital 2 (33% accepted). Common recommendations included de-escalation of antibiotics, microbiology and additional testing, medication switches, and documentation of antibiotic indications in the charts. Recommendations were communicated through face-to-face discussions, notes attached to patient medical charts, or phone calls. AMS teams also prepared a monthly summary PAF report for each intervention ward in both hospitals, and clinical pharmacists presented this report in ward meetings and attended routine clinical ward rounds.

In addition to PAF, doctors in the intervention wards also participated in evaluation activities at baseline and during the intervention period conducted by the AMS team to inform planning and monitoring, including retrospective review of antibiotic prescriptions^13^ and repeated surveys on knowledge, attitude, and practices (KAP) related to antibiotic use, AMR, and AMS; KAP surveys were also completed by doctors in the control wards (Figure 1S, Supplementary Data). Training was delivered by experts from national universities for medicine and pharmacy and hospitals for tropical diseases, focusing on antibiotic treatment for common infections, surgical prophylaxis, antibiotics, and the use of microbiological tests and interpretation of microbiology results.

### Patient and public involment

Patients and/or the public were not involved in the design, or conduct, or reporting, or dissemination plans of this research.

### Outcome measures

#### Primary outcomes

- *Antibiotic use*: The primary outcome is the amount of antibiotic use in Days of Therapy (DOT) per 1000 patient-days on a weekly time interval. Raw patient-level antibiotic prescription data from hospital information systems (HIS) were extracted together with patient administrative, diagnosis discharge outcomes and bed day information. From these data, we calculated the antibiotic use indicators overall and by Anatomical Therapeutic Chemical (ATC) classification of chemical therapeutic subgroup defined by the World Health Organization (WHO). Patients could move between study wards; each patient was counted only once under each grouping (and antibiotics used by the patient on a specific day were counted for the ward where the patient stayed on that day). We also described antibiotic use before and after the start of the AMS intervention by calculating the proportions of patients admitted to each study ward who used at least one antibiotic, and proportions of all used antibiotics by AWaRe (Access, Watch, Reserve, and Other) groups (2021 version) defined by WHO^14^.
- *Antibiotic non-susceptibility among hospital-acquired isolates:* Microbiology data were deduplicated, i.e. if a patient had several specimens collected within 30-days, then the duplicate results were excluded. Hospital-acquired isolates were defined as those from specimens sampled at least 48 hours after hospital admission (counted for the first positive sample of the same specimen type and bacterium only). Hospital-acquired isolates were identified and analysed for the change in the non-susceptibility proportion after intervention. We measured the proportions of antibiotic non-susceptibility in five common organisms identified in routine clinical investigations (specimens from all bodily sites, excluding specimens for screening purposes): *Escherichia coli, Klebsiella* spp., *Acinetobacter* spp., *Pseudomonas aeruginosa*, and *Staphylococcus aureus*. Raw data for antibiotic susceptibility testing results were extracted from the WHONET database of each hospital and interpreted using the AMR R package^15^ (interpretation using CLSI guidelines 2023). Non-susceptibility proportions were calculated as the ratio of the number of non-susceptible isolates to the number of tested isolates for a specific organism (isolates were deduplicated by patient and specimen type). We reported microbiology data following the recommendations of the MICRO framework (Supplementary Data) for the following pathogen-drug combinations which were considered relevant in the local epidemiological context:
- *E*.*coli* and *Klebsiella* spp.: third-generation cephalosporin (ceftriaxone or ceftazidime), aminoglycoside (gentamicin and one of amikacin or tobramycin), fluoroquinolone (ciprofloxacin), carbapenem (one of ertapenem, imipenem, meropenem, or doripenem);
- *P. aeruginosa* and *Acinetobacter* spp.: third-generation cephalosporin (ceftazidime), aminoglycoside (one of amikacin or tobramycin), fluoroquinolone (ciprofloxacin), carbapenem (one of imipenem, meropenem, or doripenem), piperacillin-tazobactam, aztreonam, colistin;
- *S. aureus:* Methicillin-resistant (MRSA) (oxacillin or cefoxitin).

#### Secondary outcomes

- In-hospital mortality: In-hospital mortality per 1000 admitted patients on a weekly time interval, including patients who died in hospital and those who were discharged to die at home.
- Cost of hospitalisation: Includes all costs incurred during each hospital admission as recorded in the medical record of each patient after hospital discharge. This is direct medical costs (including all types of costs: drugs, medical services, procedures, consumables, tests, bed and room services) paid to the hospital, either by the patient out of pocket or by a third-party payer (such as health insurance). Direct non-medical costs and indirect costs were not included. All costs were converted from Vietnam Dong to US Dollar in 2021 values, with costs incurred in 2019 and 2020 adjusted to the equivalent values in 2021 using Gross Domestic Product deflation rates^16^.

For antibiotic use, mortality, and hospitalisation costs, before-intevention time series were available from 1 Jan 2019 to 1 Jun 2021 in Hospital 1 and from 1 Jan 2019 to 29 Jul 2021 in Hospital 2. Antibiotic non-susceptibility data were available from 1 Jan 2014 to 31 Dec 2021 in Hospital 1, except for *S. aureus* from 1 Jan 2017 to 31 Dec 2021 and from 1 Dec 2017 to 31 Dec 2021 in Hospital 2.

### Statistical methods

All analyses were conducted separately for each hospital. The proportion of patients with any antibiotic use during their hospital stay, DOT per 1000 patient-days, DOT percentage by AWaRe classification, in-hospital mortality per 1000 admitted patients, and cost of hospitalisation were summarised by pre- and post-intervention periods for all study wards and for each ward pair. Outcome variables were visualised and decomposed to examine potential patterns, trends, and seasonality. Based on the available observations and periodic patterns, the aggregate unit, i.e. week or month, for each outcome was determined.

An interrupted time series (ITS) design was used to compare longitudinal changes in the post-intervention period to a hypothetical scenario in which the intervention did not occur. We then performed a controlled interrupted time series (CITS) analysis that incorporated both control and intervention groups into an ITS model. Segmented regression models were used to estimate the effects of the intervention for both ITS and CITS, which are shown in the supplementary data. Specifically, antibiotic non-susceptibility was modelled using logistic regression, while other outcomes were modelled using linear regressions.

Segmented regression model assumptions were checked by examining the residuals, particularly temporal correlation using ACF/PACF plots and Ljung-Box test. For linear segmented regression, an Autoregressive Integrated Moving Average (ARIMA) model was added to the regression model to adjust for non-stationarity, autocorrelation, and seasonality. ARIMA model selection was performed using an automated process in the R function auto.arima().

For logistic regression, the models were first run without any lags of antibiotic non-susceptibility proportions. In case of auto-correlation in the simulated residuals, the models were re-run afte having added lags of antibiotic non-susceptibility proportion as suggested by the ACF plots. In case of presence of overdispersion, the logistic regression models were re-run replacing the binomial distribution by a quasibinomial. Finally, for all models with less than 10 events (antibiotic use or antibiotic non-susceptibility) or non-events per co-variable, the models were re-run with L_2_ regularization (ridge) in order to avoid biases and large confidence intervals for the parameter estimates. The optimal value of the shrinkage parameter was searched by cross-validation as implemented by the cv.glmnet() function of the glmnet R package. (See additional information in the methods provided in Supplementary Data). All analyses were conducted in R (v.4.3.1; R Core Team 2022).

This study report follows the criteria described for nonrandomised evaluations of behavioral and public health interventions^17^ (see TREND Statement Checklist in Supplementary Data).

## Results

Figure 1 illustrates the inpatient admissions to the eight study wards in each hospital, both prior to and following the implementation of the AMS programme. In total, there were 45,623 patients in the intervention group and 51,444 patients in the control group in Hospital 1, while Hospital 2 reported 18,842 and 25,401 patients, respectively. A summary of patient demographic characteristics by study group, ward pair and hospital is available in Table 2S (Supplementary Data).

In Hospital 1, the most common diagnoses in the intervention group included injury/poisoning (18.3%), gastrointestinal (13.6%), infectious (12.5%), and respiratory (10.9%). In the control group, the predominant diagnoses were respiratory (29.3%), infectious (14.8%), digestive (13.4%), cardiovascular (9.8%), and genitourinary (8.0%). In Hospital 2’s intervention group, the leading diagnoses were injury/poisoning (38.3%), cardiovascular (12.2%), gastrointestinal (9.5%), and respiratory (8.2%). In the control group, the most frequent diagnoses were oncological (29.7%), gastrointestinal (26.2%), infectious (14.4%), and respiratory (9.1%) (Table 3S, Supplementary Data).

### Antibiotic use

#### Proportion of patients with any antibiotic use

Higher proportions of patients with any antibiotic use were observed in the intervention group compared to the control group across most diagnostic categories in Hospital 1, except for infectious, genito-urinary, and respiratory diagnoses during both periods. Conversely, in Hospital 2, the proportions of antibiotic use were lower for circulatory, gastrointestinal, and not-elsewhere-classified diagnoses in the intervention group (Table 3S, Supplementary Data).

The proportion of patients with any antibiotic use decreased slightly in the post-intervention period for both intervention and control group wards in Hospital 1, with respective absolute reductions of 1.3% (from 71.4% to 70.1%) and 0.6% (from 67.0% to 66.4%) (Table 1). In contrast, Hospital 2 experienced an absolute reduction in antibiotic use in intervention wards of 4.4% (from 68.3% to 63.9%) and an absolute increase of 4.3% (from 72.0% to 76.3%) in control wards (Table 1).

**Table 1.**
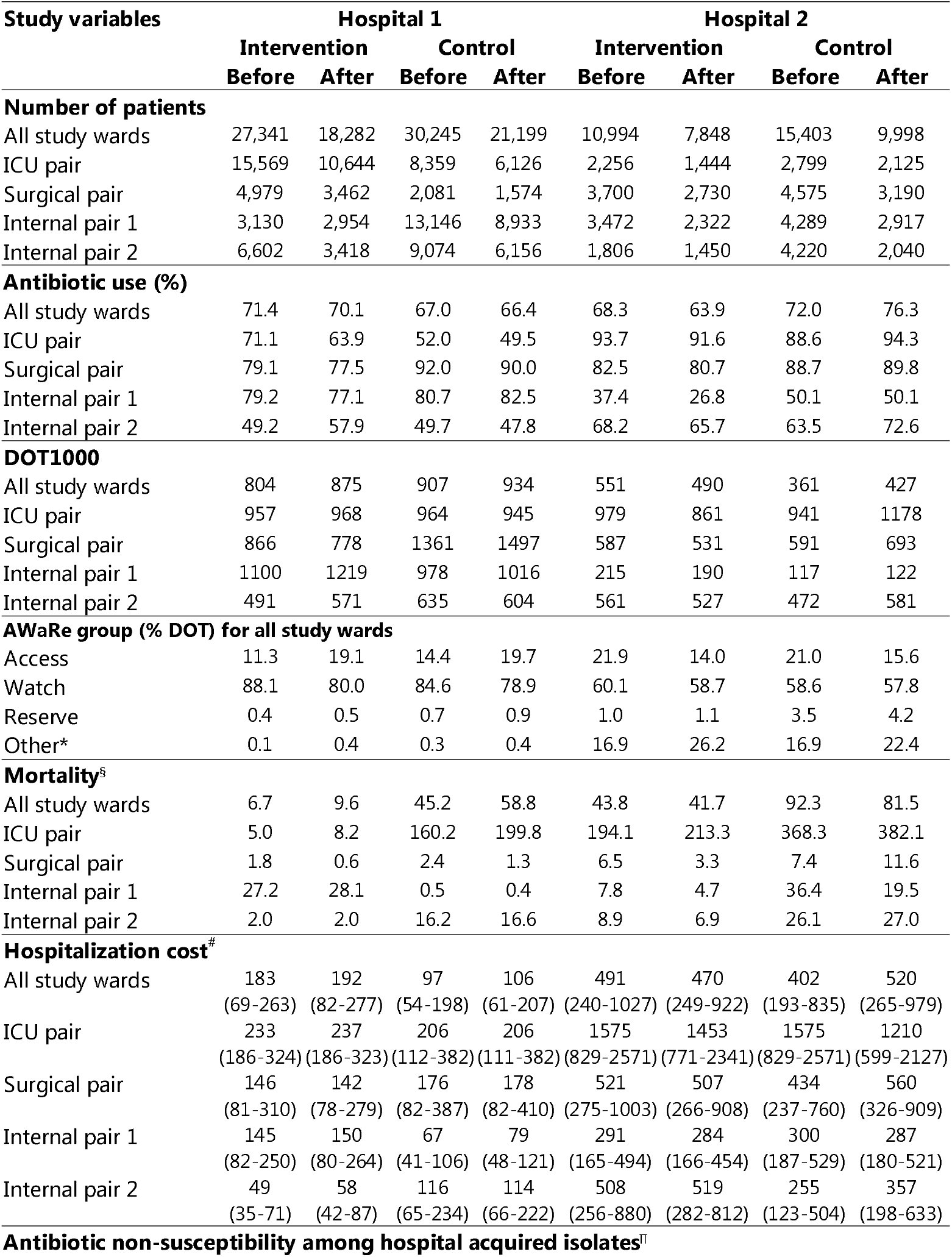
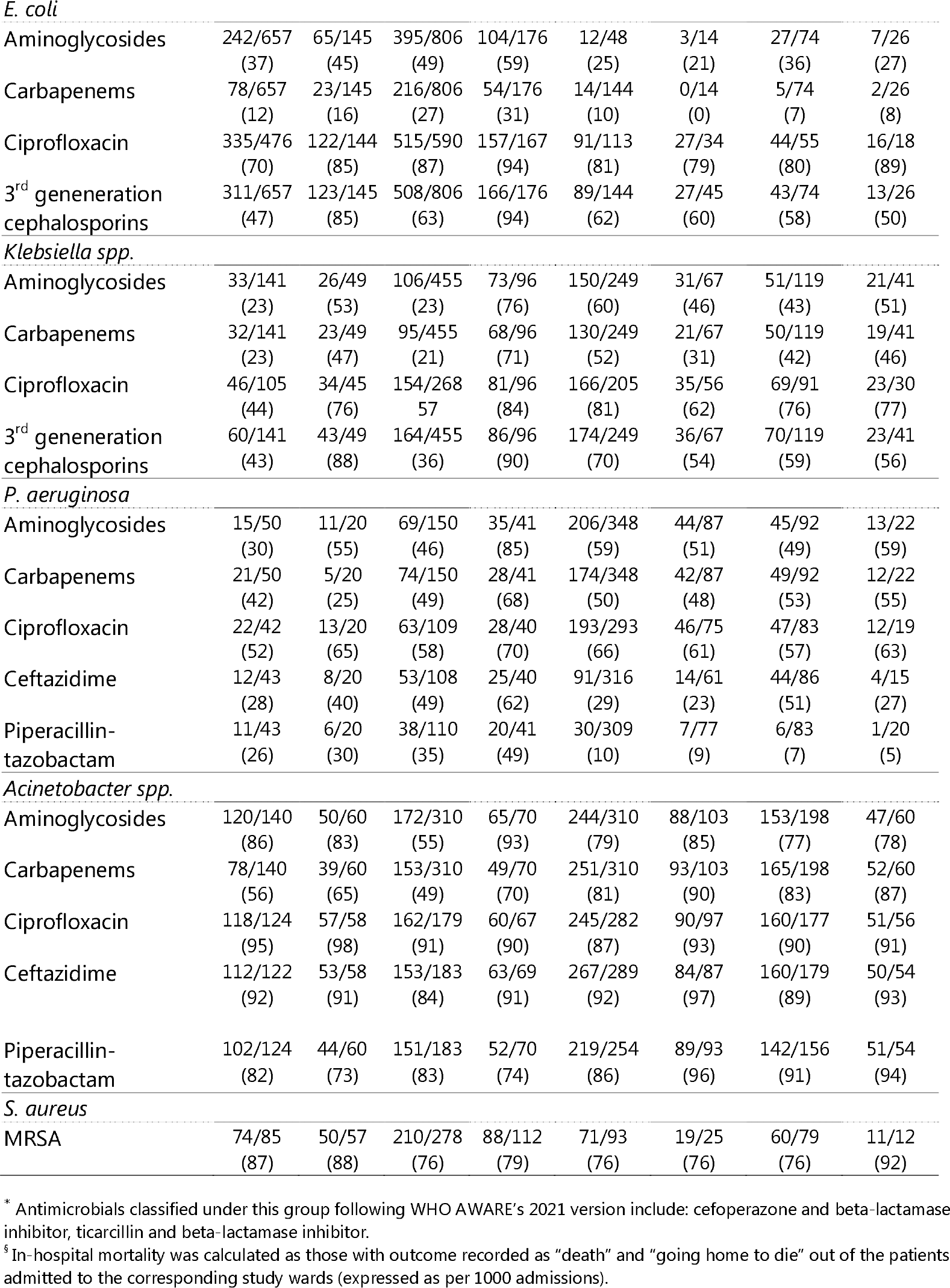
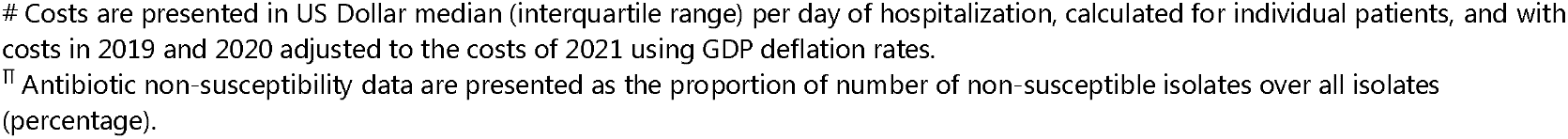
Study participants, antibiotic use, mortality, costs of hospitalization and antibiotic non-susceptibility among hospital-acquired isolates before and after the start of AMS intervention.

#### Proportion of AWaRe antibiotic classification

There was an increase in the proportion of Access antibiotics from 11.3% to 19.1% in the intervention group and from 14.4% to 19.7% in the control group at Hospital 1. Conversely, the use of Access antibiotics fell at Hospital 2, declining from 21.9% to 14.9% in the intervention group and from 21.0% to 15.6% in the control group. Additionally, there was a rise in the use of other antibiotic categories that were classified as “Not recommended” or unclassified by the WHO in the AWaRe classifications version 2021 (including cefoperazone/beta-lactamase inhibitor and ticarcillin/beta-lactamase inhibitor), increasing from 16.9% to 26.2% and from 16.9% to 22.4%, respectively (Table 1). We observed decreases in the use of certain subgroups, accompanied by increases in others, depending on the available antibiotic agents at each hospital, with more pronounced changes noted at Hospital 2 compared to Hospital 1 (Table 2S, Supplementary Data). In the intervention groups, there were decreases in the use of second-generation cephalosporins (from 13.8% to 5.2%) and glycopeptides (from 3.7% to 2.3%) at Hospital 1, and in penicillin/beta-lactamase inhibitors (from 27.9% to 24.6%), fourth-generation cephalosporins (from 5.6% to 2.5%), and fluoroquinolones (from 22.6% to 16.5%) at Hospital 2.

Based on the frequency of observations for each outcome, we determined to apply a weekly interval for antibiotic use, mortality, and hospitalisation costs, while adopting a monthly interval for antibiotic non-susceptibility, given that the number of antibiotic non-susceptible isolates is relatively low.

#### Days of antibiotic therapy per 1000 patient-days (DOT1000)

From the summary report, the overall DOT1000 increased in the post-intervention period for both the intervention and control groups in Hospital 1. In contrast, Hospital 2 experienced a decline in the post-intervention period for the intervention group, while the control group showed an increase (Table 1; for comprehensive details of numerators and denominators, refer to Table 2S, Supplementary Data).

##### ITS/CITS results for DOT1000 in the intervention group

ITS/CITS models provided evidence of consistent changes in DOT1000 for Hospital 1 regarding overall antibiotic use and certain subgroups. Specifically, the intervention was associated with an immediate (i.e. level) reduction in overall antibiotic use by 95.9 (CITS 95% CI: [10.9, 180.8]; ITS: 93.3 [0.7, 186.0]) and a reduction in the DOT1000 level of glycopeptide antibacterials by 52.7 (CITS [38.0, 67.5]; ITS: 60.2 [40.6, 79.8]). In contrast, the DOT1000 level of beta-lactamase resistant penicillins increased by 7.3 (CITS [0.2, 14.3]; ITS: 8.4 [2.7, 14.0]). The slope of the DOT1000 for second-generation cephalosporins indicates a long-term effect, with an average decrease of 1.3 (CITS [0.5, 2.0]; ITS: 1.9 [1.1, 2.8]) for each additional week, reflecting a declining long-term trend over time. Conversely, the slope of the DOT1000 for penicillin/beta-lactamase inhibitors increased by 1.9 (CITS [0.9, 2.9]; ITS: 1.6 [1.0, 2.2]). The slopes of aminoglycosides, polymyxins, and imidazole derivatives increased in ITS models by 0.4 (ITS [0.1, 0.7]), 0.1 (ITS [0.03, 0.2]), and 0.5 (ITS 95% CI: [0.1, 0.9]), respectively, while the slope of glycopeptide antibacterials DOT1000 decreased in the CITS model by 0.6 (CITS [0.1, 1.0]) (Figure 2; Figure 4S, Table 11S, Supplementary Data).

**Figure 2.**
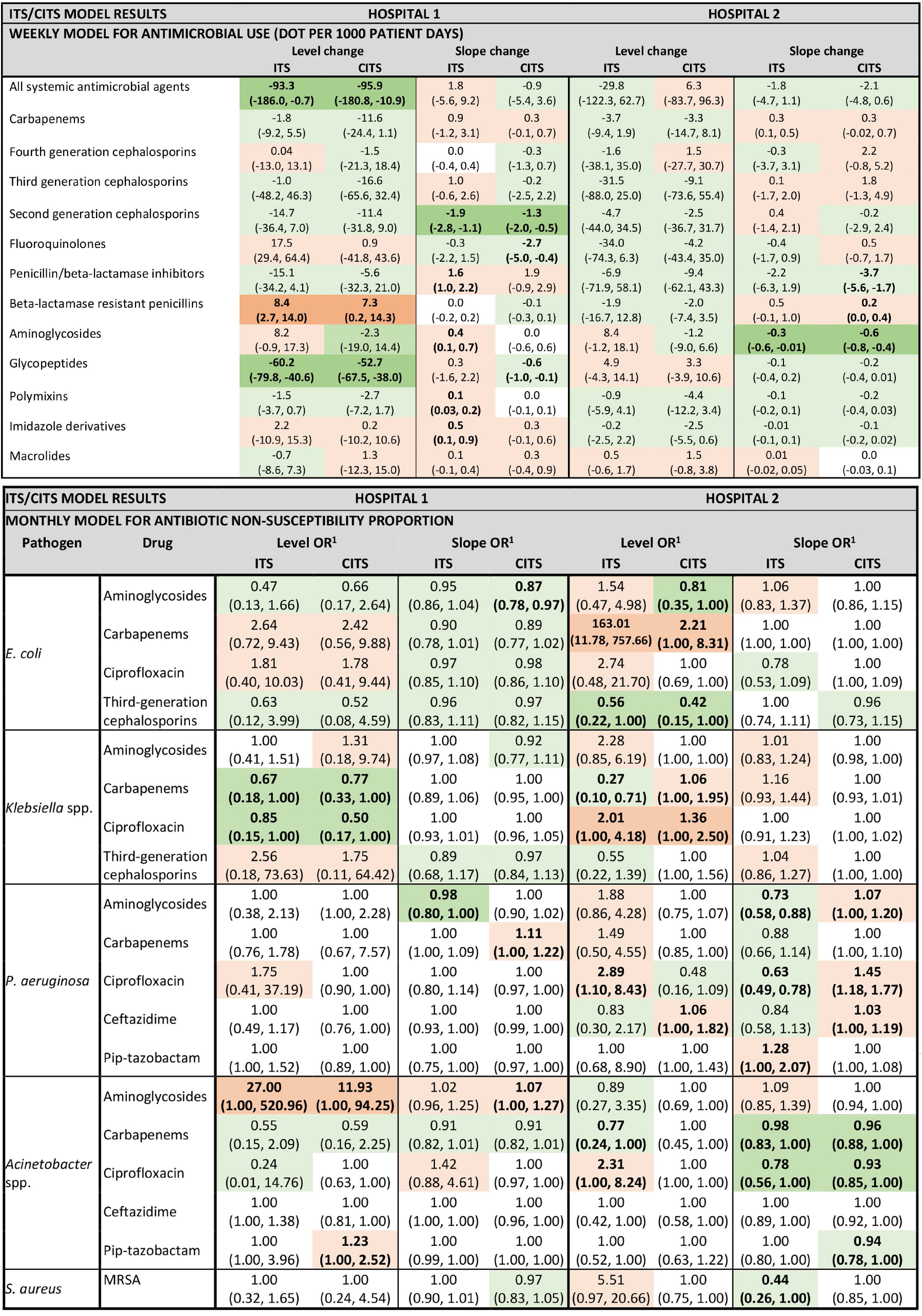
Results of ITS and CITS models for antibiotic use and antibiotic non-susceptibility among the hospital-acquired common pathogens identified from routine microbiology in the intervention group at two hospitals. ^1^ Odds ratio between non-susceptibility vs susceptibility. Bold values: statistically significant estimate; Green shades: decreasing trends; Red shades: increasing trends; Dark shades: consistent significant results for ITS/CITS models. Pip-tazobactam: Piperacillin-tazobactam.

For the ward pair subgroup analysis, the intervention was associated with a decrease in overall antibiotic DOT1000 levels in the traumatology ward by 213.0 (ITS [74.8, 351.3]), an increase in DOT1000 levels in the general medicine ward by 127.9 (ITS [6.2, 249.6]), and a decreasing slope in DOT1000 for the respiratory/musculoskeletal system ward by 3.1 (CITS [1.1, 5.0]) (Figure 5S, Table 11S, Supplementary Data).

In Hospital 2, the results from ITS and CITS were inconsistent, with some evidence of changes observed only in specific antibiotic subgroups. Notably, there was strong evidence indicating that the slope of aminoglycosides DOT1000 decreased by 0.6 (CITS [0.4, 0.8]), whereas the DOT1000 for penicillin/beta-lactamase inhibitors exhibited a significant reduction only in the CITS model by 3.7 (CITS [1.7, 5.6]), and the slope of carbapenem DOT1000 showed a significant increasing effect only in the ITS model by 0.3 (ITS [0.1, 0.5]). In the ward pair analysis, only the high-quality general medicine ward demonstrated an increase in DOT1000 by 87.6 (ITS [16.0, 159.2]) (Figure 2; Figure 5S, Table 11S, Supplementary Data).

### Antibiotic non-susceptibility

Descriptive statistics, encompassing the frequency and percentage of non-susceptibility to antibiotics among hospital-acquired isolates, as well as the intervention and control ITS/CITS models, are presented for the intervention group (Table 1; Figure 3&4). A comprehensive report of the descriptive statistics relating to other antibiotic non-susceptibility and the ITS/CITS models conducted on the control group is provided in Supplementary Data.

**Figure 3.**
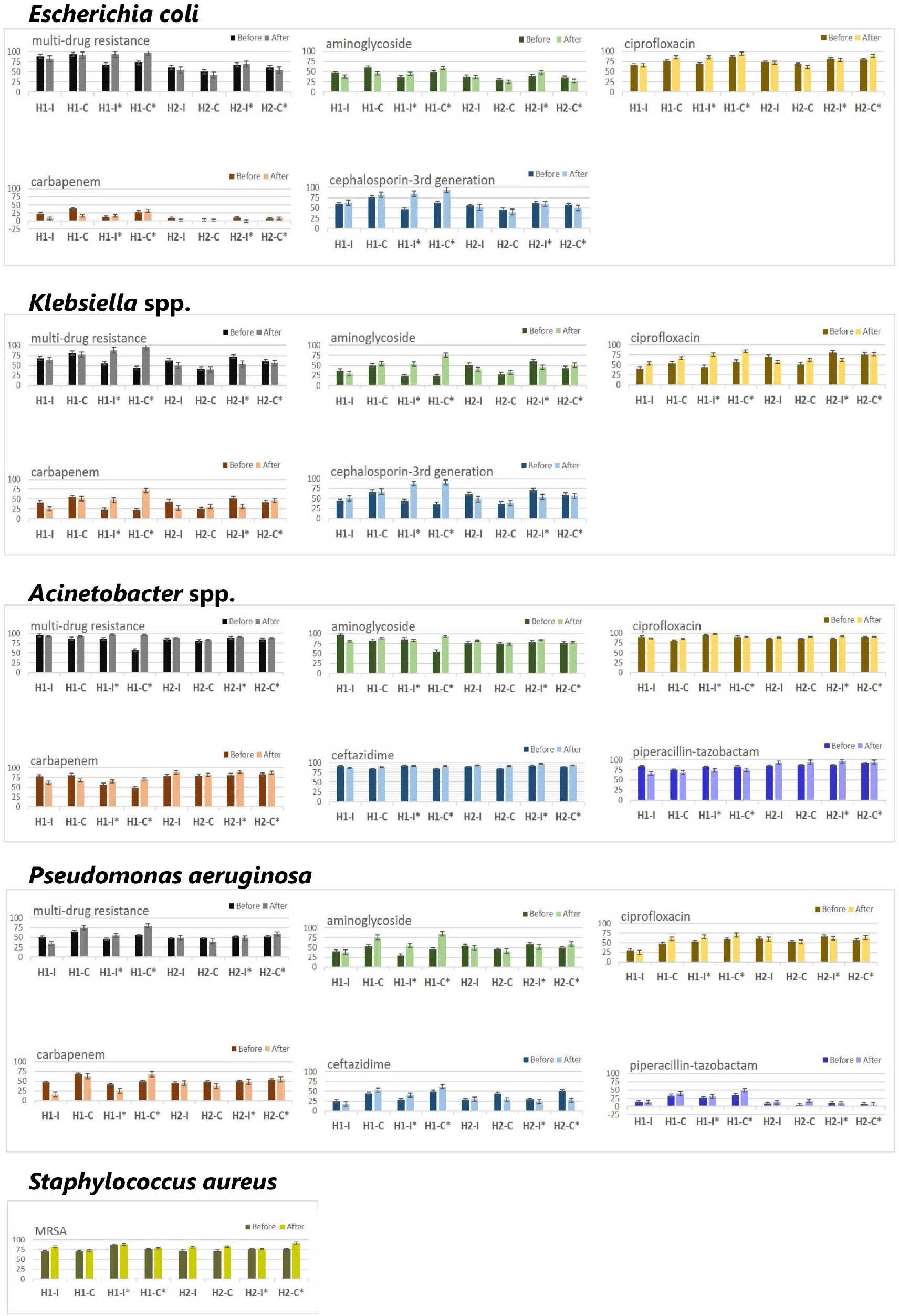
Proportion of antibiotic non-susceptibility for main pathogen-drug pairs in intervention and control groups before and after the start of AMS intervention at two hospitals; H1: Hospital 1; H2: Hospital 2; I: Intervention; C: Control; MRSA: Methicillin-resistant Staphylococcus aureus; Those columns with * present results for hospital acquired isolates, the remaining columns are for all isolates in the corresponding group.

**Figure 4.**
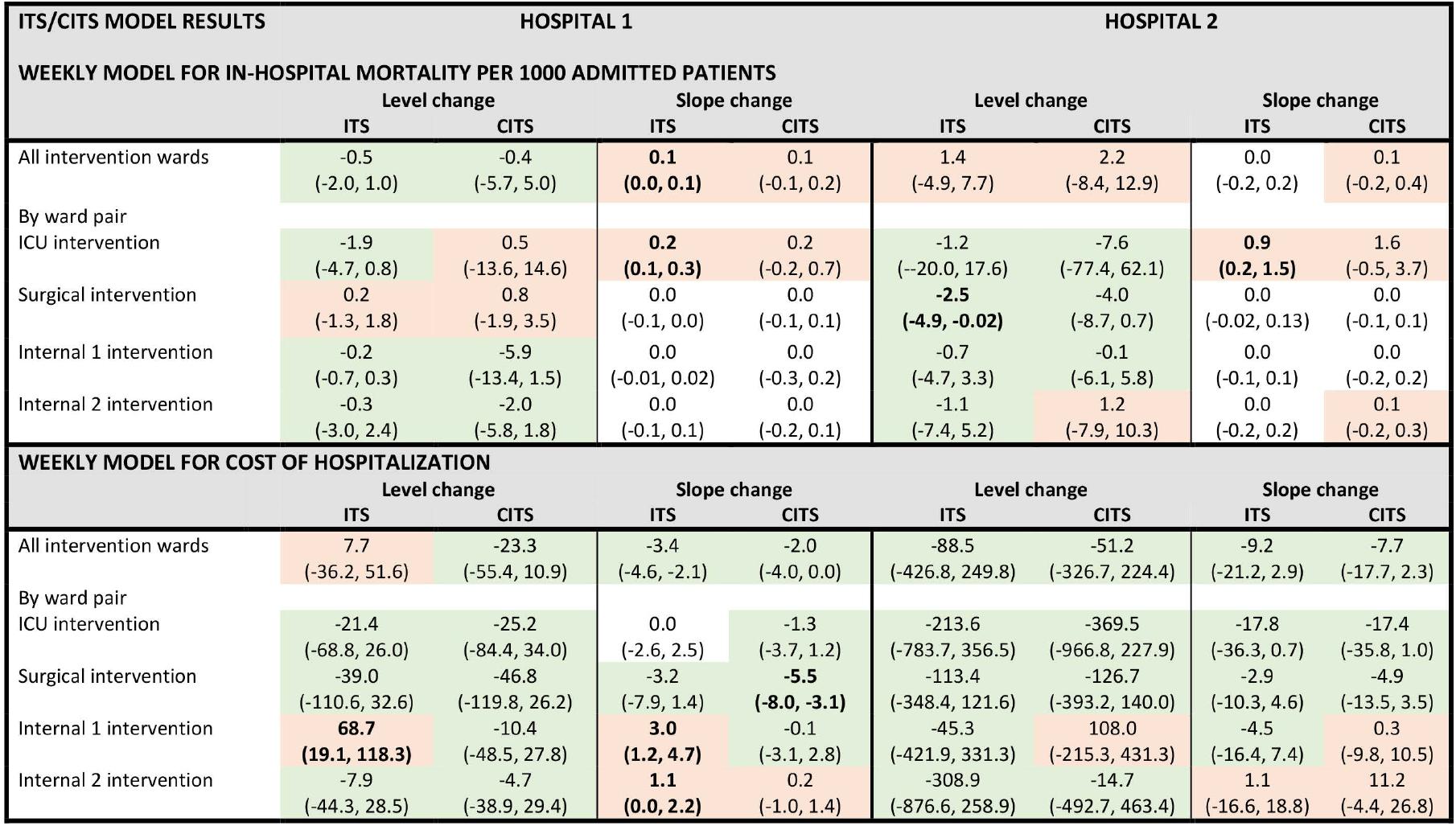
Results of ITS and CITS models for in-hospital mortality and cost of hospitalization in the intervention group at two hospitals. Bold values: statistically significant estimate; Green shades: decreasing trends; Red shades: increasing trends.

#### E. coli

Among hospital-acquired *E. coli* isolates, the percentages of non-susceptible isolates were observed to be higher during the post-intervention period in Hospital 1, whereas the opposite trend was noted in Hospital 2 (Table 1; Figure 3; Table 10S, Supplementary Data). However, in the CITS models, the AMS intervention resulted in a slope reduction for aminoglycoside non-susceptibility (odds ratio [OR] 95%CI: 0.87 [0.78, 0.97]) in Hospital 1. In Hospital 2, the level decreased for aminoglycosides (0.81 [0.35, 1.00]) and third-generation cephalosporins non-susceptibility (0.42 [0.15, 1.00]), but increased for carbapenems non-susceptibility (2.21 [1.00, 8.31]; no evidence of changes in the slopes (Figure 2; Table 12S, Supplementary Data).

#### *Klebsiella* spp

The percentages of *Klebsiella* spp. non-susceptible to antibiotics exhibited patterns similar to those observed in *E. coli*, characterised by a higher percentage of non-susceptible isolates in the post-intervention phase for Hospital 1, and a lower percentage for Hospital 2 (see Table 1; Figure 3; Table 10S, Supplementary Data). Analysis using ITS/CITS models showed consistent changes only in the levels of non-susceptibility, with a decrease in the level of non-susceptibility to carbapenems (CITS 0.77 [0.33, 1.00]) and ciprofloxacin (0.50 [0.17, 1.00]) in Hospital 1. Conversely, in Hospital 2, the intervention was associated with a decreased level of non-susceptibility to carbapenems in the ITS model (ITS 0.27[0.10, 0.71]) but increased in the CITS model (1.06 [1.00, 1.95]), and increased in both models for ciprofloxacin (ITS 2.01 [1.00, 4.18], CITS 1.36 [1.00, 2.50]) (Figure 2; Table 12S, Supplementary Data).

#### P. aeruginosa

Similar trends were observed in the percentages of non-susceptible *P. aeruginosa* isolates across both hospitals (Table 1; Figure 3; Table 10S, Supplementary Data). In Hospital 1, the results from the ITS/CITS analyses indicated an increase in the slope of non-susceptibility to carbapenems in the CITS model (1.11 [1.00, 1.22]), and a decrease for aminoglycosides but in the ITS model only (0.98 [0.80, 1.00]). Conversely, Hospital 2 exhibited mixed results between both models in the slopes of non-susceptibility to ceftazidime (CITS 1.03 [1.00, 1.19]; ITS 0.84 [0.58, 1.13]), aminoglycosides (CITS 1.07 [1.00, 1.20]; ITS: 0.73 [0.58, 0.88]), and ciprofloxacin (CITS 1.45 [1.18, 1.77]; ITS: 0.63 [0.49, 0.78]) (Figure 2; Table 12S, Supplementary Data).

#### *Acinetobacter* spp

The percentages of non-susceptibility showed varying patterns in the descriptive analysis. Notably, the non-susceptibility percentages were heterogeneous in Hospital 1, whereas all antibiotic non-susceptibility percentages were elevated in Hospital 2 (Table 1; Figure 3; Table 10S, Supplementary Data). The CITS models indicated an increase in the level (11.93 [1.00, 94.25] and slope of non-susceptibility to aminoglycosides (1.07 [1.00, 1.27]), and in the level of non-susceptibility to piperacillin tazobactam (1.23 [1.00, 2.52]) in Hospital 1. In contrast, a reduction was noted in the slope of non-susceptibility to carbapenems (CITS 0.96 [0.88, 1.00]), ciprofloxacin (CITS 0.93 [0.85, 1.00]) and piperacillin tazobactam (CITS 0.94 [0.78, 1.00]) in Hospital 2 (Figure 2; Table 12S, Supplementary Data).

#### MRSA

The percentages of MRSA remained stable during the post-intervention period in both hospitals (Table 1; Figure 3; Table 10S&12S, Supplementary Data), except for the decreasing slope only observed in the ITS model of Hospital 2 (0.44 [0.26, 1.00]) (Figure 2; Table 12S, Supplementary Data).

### In-hospital mortality and costs

There was a non-significant decrease in the level of mortality with increasing slope in Hospital 1 intervention wards overall, while increasing levels and slopes were observed in Hospital 2 (Table 2S; Table 13S, Supplementary Data). The ITS/CITS models for mortality show inconsistent results for the specific wards of both hospitals (Figure 4; Table 13S, Supplementary Data), with insignificant changes reported in all CITS models.

Considering the costs for monthly activities of the AMS team (coordinator/pharmacists) and on-site training as regular activities as part of the AMS programme at each hospital, the one-year costs for AMS implementation were approximately 2,809 USD for Hospital 1 and 2,283 USD for Hospital 2 (Table 1S, Supplementary Data).

Regarding hospitalization costs, in both ITS and CITS models, we observed consistent decreasing levels and slopes overall and across most study wards in both hospitals. Particularly, the cost of hospitalisation in the intervention surgical ward decreased in slope by 5.5 (CITS [3.1, 8.0]) US Dollar in Hospital 1 (Figure 4; Table 14S, Supplementary Data).

## Discussion

We examined the impact of a PAF intervention on antibiotic use and antibiotic non-susceptibility for multiple pathogen-drug pairs among common hospital-acquired bacterial isolates. This approach acknowledged the potential “squeezing the balloon” phenomenon in AMS programmes^18^, wherein limiting the use of specific antibiotics may lead to counteracting and unintended changes in the use of other antibiotics and drug resistance mechanisms. Our findings support this assumption, with observed decreases in some antibiotic groups but increases in others, reflecting the complexity of antibiotic use and resistance in provincial hospitals with high antibiotic consumption.

The observed impact models varied between the two hospitals, reflecting the context-specific nature of the implementation. In Hospital 1, ITS and CITS models provide evidence that overall antibiotic consumption immediately decreased in the intervention group with consistent immediate and long-term impacts on antibiotic non-susceptibility for hospital-acquired *E. coli*. In contrast, the impact of AMS on antibiotic use was more limited in Hospital 2, with some evidence of positive long-term effects on non-susceptibility of *Acinetobacter* spp. These heterogeneous findings and the lack of significant intervention effects in various subgroups or outcome indicators were likely influenced by confounders, such as differences in community antibiotic pressure, patient characteristics, baseline resistance profiles, resource availability (number of clinical pharmacists dedicated to the PAF activity), hospital culture and staff engagement, and the behaviour and collaboration of doctors and pharmacists towards the AMS intervention, which could have impacted both the intervention and the antibiotic use outcomes. Furthermore, the effects on antibiotic consumption likely contributed to the inconsistent impacts on antibiotic non-susceptibility.

One of the inclusion criteria for selecting the wards was the willingness of the ward head to participate. This may have helped increase the collaboration of doctors in the intervention wards and their compliance with the recommendations of the AMS team in the PAF activity, and thus possibly limits the generalizability of the study. However, the PAF activity reached only a small number of patients during the implementation period (1,890/45,623 or 4.1% in Hospital 1, and 1,628/18,842 or 8.6% in Hospital 2). This suggests the change in practice following the PAF activity is likely to be from the systems level rather than the individual patient level, particularly the impact on reducing overall antibiotic use in Hospital 1. The presence of clinical pharmacists on the clinical wards to regularly audit antibiotic prescriptions may have increased the compliance of doctors to prescription guidelines in general and in the surgical ward (traumatology) in particular, potentially providing a generalizable evidence for the impact of this non-restrictive AMS intervention for other settings.

For mortality, the results of our CITS models indicate that AMS implementation at the hospital-wide level could improve antibiotic use without causing negative consequences on patient outcomes. The increasing trends in mortality were found in ITS models for both intervention and control ICUs, but not in the CITS models, highlighting the importance of using a control ^19^ when evaluating impact of interventions. Further examination revealed increased post-intervention mortality for most diagnoses among ICU patients in both hospitals, particularly for diseases of the respiratory system and abnormal clinical and laboratory findings (Table 15S, Supplementary Data). Nonetheless, further study is needed to fully investigate the impact of AMS in ICU patients, considering the differential effects that might happen to patients of different diagnoses. The limited number of data points in our current datasets does not provide sufficient power to identify the changes in specific clinical diagnoses over time in ICUs.

Generally, AMS programmes in Vietnamese hospitals must follow national guidelines, which require forming an AMS committee, assigning roles, developing hospital-specific policies, and implementing restricted antibiotic lists^10 20^. Most hospitals responded to the national guidelines by quickly convening committees, while specific actions and interventions for monitoring and improving antibiotic use and resistance remained limited due to poor leadership commitment, lack of dedicated staff, and weak IT capacity and lab resources^8^. Many hospitals established pre-authorization systems for restricted antibiotics to ensure compliance with national guidelines, often tied to social health insurance reimbursement, particularly for expensive and potentially overused medical services^21^. In this context, clinical pharmacist-led audits of antibiotic prescriptions, with constructive feedback to doctors, offer a proactive intervention to enhance collaboration among healthcare professionals. Our feasibility study showed that this approach could be integrated into routine clinical pharmacy work^9^. The intervention involved reviewing active antibiotic prescriptions during pharmacist visits, providing recommendations for inappropriate practices, and assessing changes in total antibiotic use and specific groups. We measured changes in both total antibiotic use and specific groups to assess the intervention’s impact. Increased collaboration among prescribing doctors, clinical pharmacists, microbiologists, and infectious disease specialists helped optimise antibiotic use for individual patients^18^.

Literature supports the causal relationships between antibiotic use in hospitals and resistance prevalence among hospital-acquired isolates antibiotics^22^. Mixed results from ITS and CITS models in our study suggest the presence of history bias and other events impacting the control group^19^. Previous ITS studies, often lacking control groups, reported mixed results as shown in a systematic review of studies by 2018^5^ as well as in more recent studies in the US^23–26^, Spain^27–29^, Germany^30^, Japan^31^, Brazil^32^, Korea^33^, and China^34^. To our knowledge, only one observational study in a 627-bed hospital in Canada used community-acquired isolates as a control time series^6^. This study demonstrated AMS impact in reducing the incidence of hospital-acquired multidrug-resistant organisms by 12.6%. However, this approach could not account for the concurrent interventions, such as IPC or other programmes within hospital settings, that could affect hospital-acquired resistance.

The significant strength of our study is the inclusion of a control group, allowing to account for concurrent events, such as the COVID-19 pandemic and IPC measures. In particular, AMS activities and staff attention to optimal antibiotic prescribing may have been negatively affected in the last few weeks of the intervention period in Hospital 1 due to the early effects of the fourth wave of the COVID-19 pandemic in Vietnam in April – May 2021^12^. We assumed such non-intervention events had broadly similar effects on both the intervention and control groups. Along with the strengths of employing an implementation research design as reported previously^9^, we demonstrated the practicality of using a participatory action research approach to evaluate a healthcare intervention that could not be ethically assessed through traditional randomised controlled trials^35^. We also showed that behaviour change interventions, such as prospective audit and feedback, can be safely integrated into routine clinical practice without causing negative consequences on patient outcomes in resource-limited settings like Vietnam, under existing national leadership and guidance.

Limitations include the relatively short timeframe for the pre- and post-intervention periods, which restricted our ability to fully capture seasonal and historical trends in antibiotic use, especially when there were frequent shortages and stock-outs of antibiotics, as well as changes in drug bidding cycles and health insurance policies that could affect prescribing practices in each hospital. There was an increasing trend in the use of fixed-dose combinations in Hospital 2 which could raise concerns about substitution effects as a result of the AMS intervention. Future studies with longer pre- and post-intervention periods will help investigate in-depth these specific changes over time to inform the design of interventions. Despite efforts to extract data for a longer pre-intervention period, we were unable to use data before 2019 for analysing antibiotic and clinical outcomes in the control and intervention groups due to changes in the HIS. However, since these system-level changes are likely to have had similar effects on both the intervention and control groups, we expect the estimates under the controlled analyses (CITS models) to be relatively robust to such system-level changes. Longer time series for antibiotic non-susceptibility data were available from laboratory systems, though missing patient identifiers impacted model performance. Nevertheless, our analysis used more data points than the minimum suggested for interrupted time-series from a simulation-based power calculation^36^. Another limitation, intrinsic to our implementation study design, is that there can be a “spillover effect”, i.e. an unintended impact of the intervention on the prescribing practices of the control wards. This may have diluted our estimated impact of the intervention on antibiotic use in the intervention group in the CITS models.

In addition, microbiology data quality (which could be affected by ward-specific variations in specimen collection practices and testing frequency), the absence of molecular data on resistance mechanisms and potential misclassification of hospital-acquired infections limits our ability to link antibiotic use changes to resistance dynamics. Finally, we were not able to collect more granular data for cost analyses as only the total cost of hospitalization was available in the cost data extracted from HIS. In order to collect more detailed costs, we are now conducting an additional study on a specific patient population and will use these data to inform our upcoming cost analyses for the AMS programme.

Our study provides initial evidence on the complex effects of AMS interventions on antibiotic use and resistance, and on how quickly changes in antibiotic use lead to reverse resistance in a particular organism^37^. These findings could be generalised to similar settings with limited resources in Asia. A combination of strategies, including drug discovery, resistance monitoring and novel interventions, is necessary to respond to current resistance phenotypes and to anticipate the evolution of antibiotic resistance in hospital settings. This is particularly challenging for hospitals in low- and middle-income countries, which have limited resources, where infectious diseases are prevalent, antibiotic use is high, and environmental reservoirs accelerate resistance spread. Surveillance data on the mechanisms of emergence and transmission of antibiotic resistance within and between hospital reservoirs, along with extended evaluation periods, are needed to further clarify the impact of AMS interventions and antibiotic use, and to inform more effective, large-scale control and policy measures across hospitals^38^.

In conclusion, this study highlights the interdependent changes in antibiotic use and antibiotic resistance driven by AMS programmes, employing pharmacist-led prospective review of antibiotic prescriptions and feedback to doctors in provincial-level hospitals in Vietnam. Our findings confirm that developing and strengthening the surveillance of antibiotic resistance, alongside AMS implementation and IPC measures, is essential to monitor and respond to resistance dynamics in hospitals effectively. These efforts will help to ensure that interventions keep pace with the rapid evolution of antibiotic resistance.

## Data Availability

All data produced in the present study are available upon reasonable request to the authors.

## Acknowledgements

We acknowledge the support and collaboration of the Medical Services Administration of the Viet Nam Ministry of Health and the World Health Organization Office in Viet Nam during the planning of this study.

## Contributors

VTLH, HRvD, EDA and DJA obtained funding and contributed to all aspects of the study design. VTLH and HRvD had overall responsibility for the study. LMQ, NTTH, VHV, CMD, VTHDE, PNT supervised and coordinated the running of the study with support from EDA, NTCT, TAQ, LNMH and NHK. VTLH, LQT and VTTD analysed the data with supervision and input from MC, BC, TK, and HCT. VTLH was responsible for the drafting of the manuscript. All authors gave approval for the final version of the manuscript.

## Patient consent for publication

Not required for this research.

## Ethics approval

The study protocol was approved by the Oxford University Tropical Research Ethics Committee (OxTREC Reference 526-19), and the Ethics Committee of National Hospital of Tropical Diseases (08/HĐĐĐ-NĐT 31 May 2019). The conduct of this study conformed to the principles embodied in the Declaration of Helsinki.

## Data availability

De-identified data may be obtained from the hospitals participating in this study when a data sharing agreement is in place.

